# A systematic review and meta-analysis on the effectiveness of an invasive strategy compared to a conservative approach in elderly patients with non-ST elevation acute coronary syndrome

**DOI:** 10.1101/19004044

**Authors:** Joan Dymphna P. Reaño, Maria Grethel C. Dimalala, Louie Alfred B. Shiu, Karen V. Miralles, Noemi S. Pestaño, Felix Eduardo R. Punzalan, Bernadette Tumanan-Mendoza, Michael Joseph T. Reyes, Rafael R. Castillo

**Affiliations:** Fellow in Adult Cardiology, Manila Doctors Hospital, Manila, Philippines; Fellow in Interventional Cardiology, Manila Doctors Hospital, Manila, Philippines; Consultant in Adult Cardiology, Manila Doctors Hospital, Manila, Philippines; Consultant in Interventional Cardiology, Manila Doctors Hospital, Manila, Philippines; Professor in Cardiovascular Medicine, Adventist University of the Philippines, Silang, Philippines; Dean Emeritus, FAME Leaders Academy, Makati, Philippines

**Keywords:** Elderly, non-ST Elevation myocardial infarction, acute coronary syndrome, invasive strategy, conservative treatment, coronary artery disease, ACS MACE, CVD in Philippines

## Abstract

**Background:** Elderly patients, 65 years old and older, largely represent (>50 %) of hospital- admitted patients with acute coronary syndrome (ACS). Data are conflicting comparing efficacy of early routine invasive (within 48-72 hours of initial evaluation) versus conservative management of ACS in this population.

**Objective:** We aimed to determine the effectiveness of routine early invasive strategy compared to conservative treatment in reducing major adverse cardiovascular events in elderly patients with non-ST elevation (NSTE) ACS.

**Data Sources:** We conducted a systematic review of randomized controlled trials through PubMed, Cochrane, and Google Scholar database.

**Study Selection:** The studies included were randomized controlled trials that evaluated the effectiveness of invasive strategy compared to conservative treatment among elderly patients ≥ 65 years old diagnosed with NSTEACS. Studies were included if they assessed any of the following outcomes of death, cardiovascular mortality, myocardial infarction (MI), stroke, recurrent angina, and need for revascularization. Five articles were subsequently included in the meta-analysis.

**Data Extraction:** Three independent reviewers extracted the data of interest from the articles using a standardized data collection form that included study quality indicators. Disparity in assessment was settled by an independent adjudicator.

**Data Synthesis:** All pooled analyses were based on fixed effects model. A total of 2,495 patients were included, 1337 in the invasive strategy group, and 1158 in the conservative treatment group.

**Results:** Meta-analysis showed less incidence of revascularization in the invasive (2%) over conservative treatment groups (8%), with overall risk ratio of 0.31 (95% CI 0.16-0.61, I^2^ =0%). There was also less incidence of stroke in the invasive (2%) versus conservative group (3%) but this was not statistically significant. A significant benefit was noted in the reduction of all-cause mortality (RR 0.63, 95% CI 0.55-0.72, I^2^=84%) and myocardial infarction (RR 0.62, 95% CI 0.49- 0.79, I^2^=63%) but with significant heterogeneity.

**Conclusion:** There was a significantly lower rate of revascularization in the invasive strategy group compared to the conservative treatment group. In the reduction of all-cause mortality and MI, there was benefit favoring invasive strategy but with significant heterogeneity. These findings do not support the bias against early routine invasive intervention in the elderly group with NSTEACS. However, further studies focusing on the elderly with larger population sizes are still needed.

## I. INTRODUCTION

Based on the World Health Organization’s Global Burden of Disease report, ischemic heart disease (IHD) is the overall leading cause of death worldwide.^1^ Although the annual number of hospital discharges for acute coronary syndromes (ACS) in developed countries has declined slowly over the past two decades, the number has increased in developing countries.^2^ In the Philippines, cardiovascular disease (CVD) remains the leading cause of mortality.^3^ The Philippine Heart Association ACS registry reported that ACS is prevalent in the age range 51-70, with mean age group of 66 years old. ^3^

The most recent American College of Cardiology/American Heart Association (ACC/AHA 2014) and the European Society of Cardiology (ESC 2015) guidelines for non–ST segment elevation ACS (NSTEACS) reflect medical advancements in therapeutics and strategies of care leading to improved survival in ACS, but this was mainly observed in relatively younger individuals (<65 years of age) and in men. These guidelines emphasize intensive and early medical and interventional therapy, particularly for those at high risk.^4,5,6^

The 2014 AHA/ACC NSTEACS Guidelines generally recommend that older patients with NSTEACS should be treated with goal-directed medical therapy, together with an early invasive strategy, and revascularization as appropriate.^5^ The 2015 ESC Guidelines for the Management of ACS, on the other hand, recommend that decisions on elderly patients with NSTEACS should be based on ischemic and bleeding risks, estimated life expectancy, comorbidities, quality of life, patient values and preferences, and the estimated risks and benefits of revascularization. ^6^ Despite the guidelines, older patients are less likely to undergo procedures after an NSTEACS than younger patients due in part to patient and practitioner concerns about the increased risk of complications.^7,8,9^

Due to conflicting results of studies, lack of specific recommendations from the abovementioned guidelines, and the paucity of data on early invasive strategy versus conservative treatment for NSTEACS in elderly patients, this meta-analysis was conducted to focus on this special population to compare benefits and risks of early invasive therapy versus conservative management.

## II. RESEARCH QUESTION

Among elderly patients aged ≥ 65 years old with NSTEACS, how effective is invasive strategy compared to conservative treatment in preventing major adverse cardiovascular events (MACE)?

## III. OBJECTIVES

### General

To determine the effectiveness of invasive strategy compared to conservative treatment in reducing MACE among elderly patients with NSTEACS.

### Specific

Among elderly patients with NSTEACS, to determine the effectiveness of invasive strategy compared to conservative treatment, in 6 months (short-term) to 3 years (long-term), in reducing:

a. Death or all-cause mortality;
b. Cardiovascular mortality;
c. Myocardial infarction (MI);
d. Stroke;
e. Recurrent angina;
f. Need for revascularization.

## IV. METHODOLOGY

### Study Registration

Prior to the conduct of the research, the study was registered and approved by the Committee on Research (CORES) of Manila Doctors Hospital.

### Criteria for considering studies for this review

The studies included were randomized controlled trials that evaluated the effectiveness of invasive strategy compared to conservative treatment among elderly patients ≥ 65 years old diagnosed with NSTEACS. Studies were included if any of the outcomes assessed were: death, cardiovascular mortality, MI, stroke, recurrent angina, and need for revascularization.

### Definition of terms

1. **Invasive strategy or early invasive strategy** –Routine early (within 48-72 hours of initial evaluation) cardiac catheterization, followed by PCI, CABG, or continuing medical therapy, depending on the coronary anatomy.
2. **Conservative treatment** - Initial optimal medical management, with cardiac catheterization reserved for patients with recurrent ischemia at rest or after a non-invasive stress test, followed by revascularization if the anatomy is suitable.
3. **Elderly patients** – Patients aged 65 years or older (WHO, 2000), with or without comorbidities.
4. **Non-ST elevation acute coronary syndrome (NSTEACS)** – Unstable angina, with or without ST segment depression on electrocardiogram with normal or raised blood concentration of troponin T or I. Elevated troponin was defined as a value exceeding the 99th percentile of a normal population at the local laboratory at each participating site.

### Search methods for identification of studies

Systematic computerized search (APPENDIX A) was performed using the Pubmed and Cochrane databases. MESH and free text of the following main key terms were used: “randomized controlled trials”, “elderly”, “non-ST elevation acute coronary syndrome”, “invasive strategy”, “conservative management”, “invasive strategy versus conservative strategy”, “major adverse cardiovascular events”, “all-cause mortality”, “cardiovascular mortality”, “myocardial infarction”, “stroke”, “recurrent angina”, “need for revascularization”. The last search was done on 10 August 2017.

Eligibility assessment was performed independently in an unblinded standard manner by three reviewers. The literature search identified 322 possible articles. Of these, 69 were relevant, particularly they involved studies related to ACS. Prospective cohort studies and post hoc analyses were excluded. Of the 69 articles, 55 were excluded due to different intervention since they did not involve comparing invasive versus conservative management in ACS. After assessing 14 articles for eligibility, 8 articles with different population and methods were excluded (details for the titles of the studies and reasons for exclusion are listed in APPENDIX D). One article was possibly eligible but did not report the event rates per treatment group. To access needed data in this particular study, correspondence with the author via email was done, but with no reply from the author until the time of writing. Five articles were subsequently included in the meta-analysis (Figure 1).

**Figure 1.**
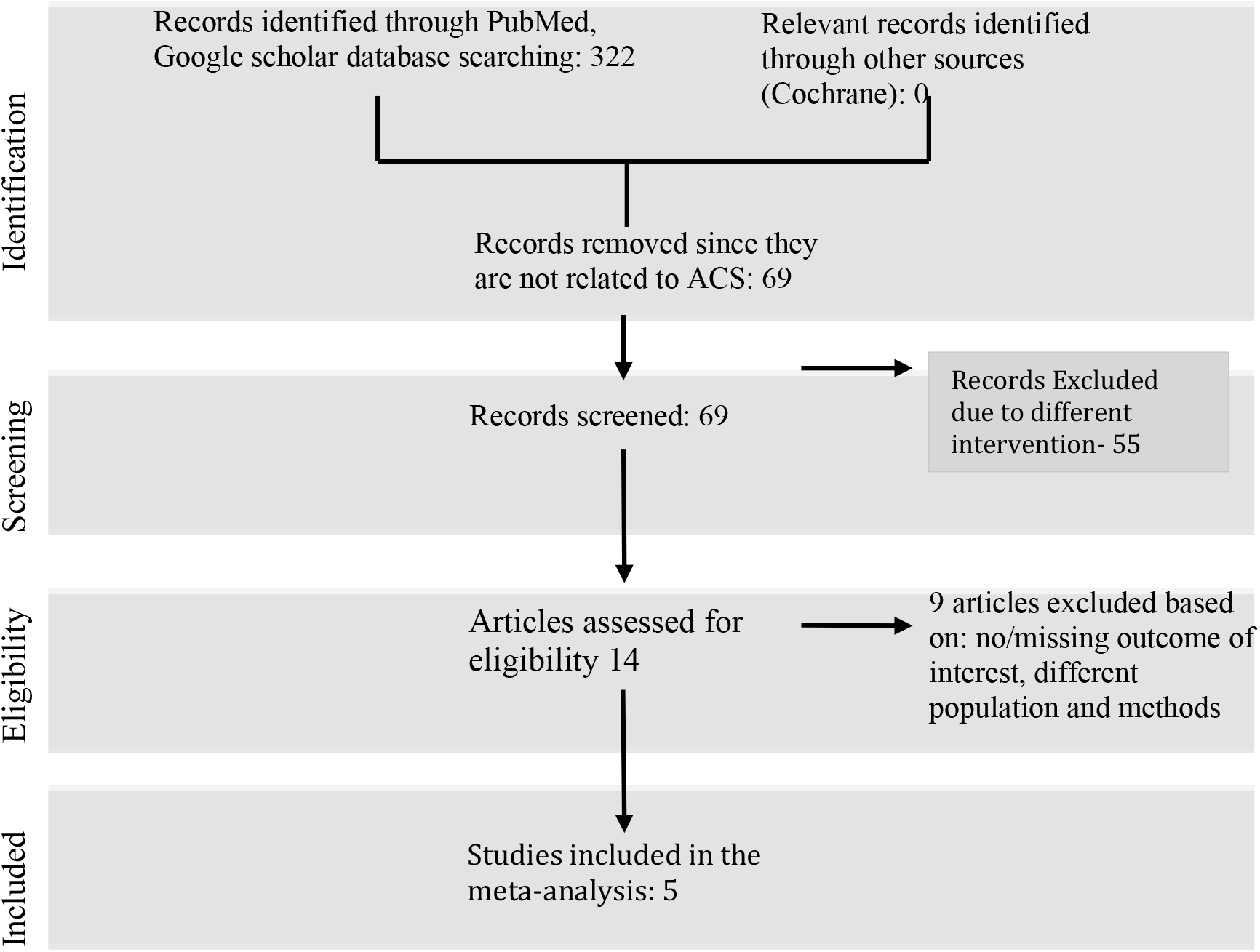
Search strategy for identification of studies.

### Assessment of risk bias of included trials

Three independent reviewers extracted the data of interest using a standardized data collection form and individually appraised each trial. The reviewers discussed the quality of included trials, outcomes to be collected, and risks of bias. Disparity in assessment was settled by an independent adjudicator. The assessment of random sequence generation, allocation concealment, incomplete outcome data, blinding of participants and personnel, blinding of outcome assessment, and intention-to-treat analysis was done using the quality scale for meta-analytic review, the Cochrane Collaboration Tool for Risk of Bias.

### Data analysis

Review Manager 5.3 was used to analyze the data. Analysis of dichotomous data was done using risk ratio, 95% confidence interval, and Mantel-Haenszel method with fixed effects model. Heterogeneity between trials was tested using a standard Chi-square test and I^2^ statistics. The p- value of <0.10 was considered to be statistically significant and I^2^ of ≥50% is considered to have high heterogeneity.

### Description of studies

Five randomized controlled trials involving a total of 2,495 patients met the inclusion criteria. The data on population characteristics, intervention type, and measured outcomes were extracted from each trial (Table 1). Four of the trials included elderly patients with NSTEACS aged ≥ 70 years while one trial included patients ≥ 65 years old.^10^ The studies compared the effectiveness of early invasive strategy (treatment group) versus optimum medical treatment (control group) in the management of NSTEACS in elderly patients.

**Table 1.**
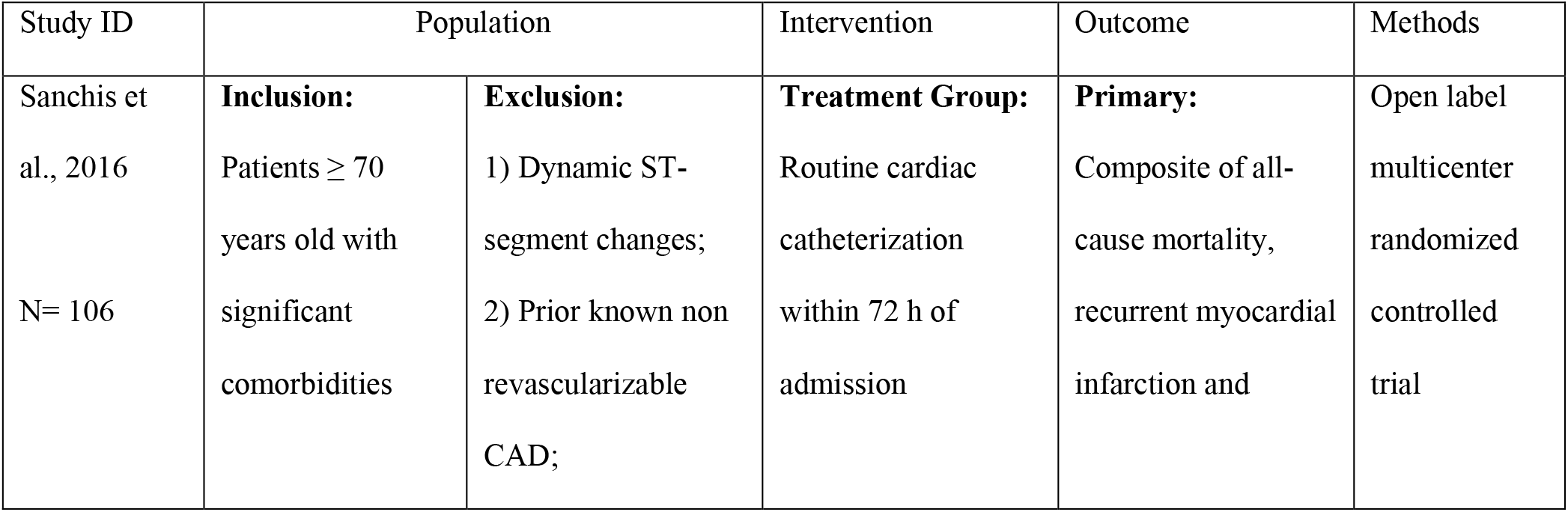
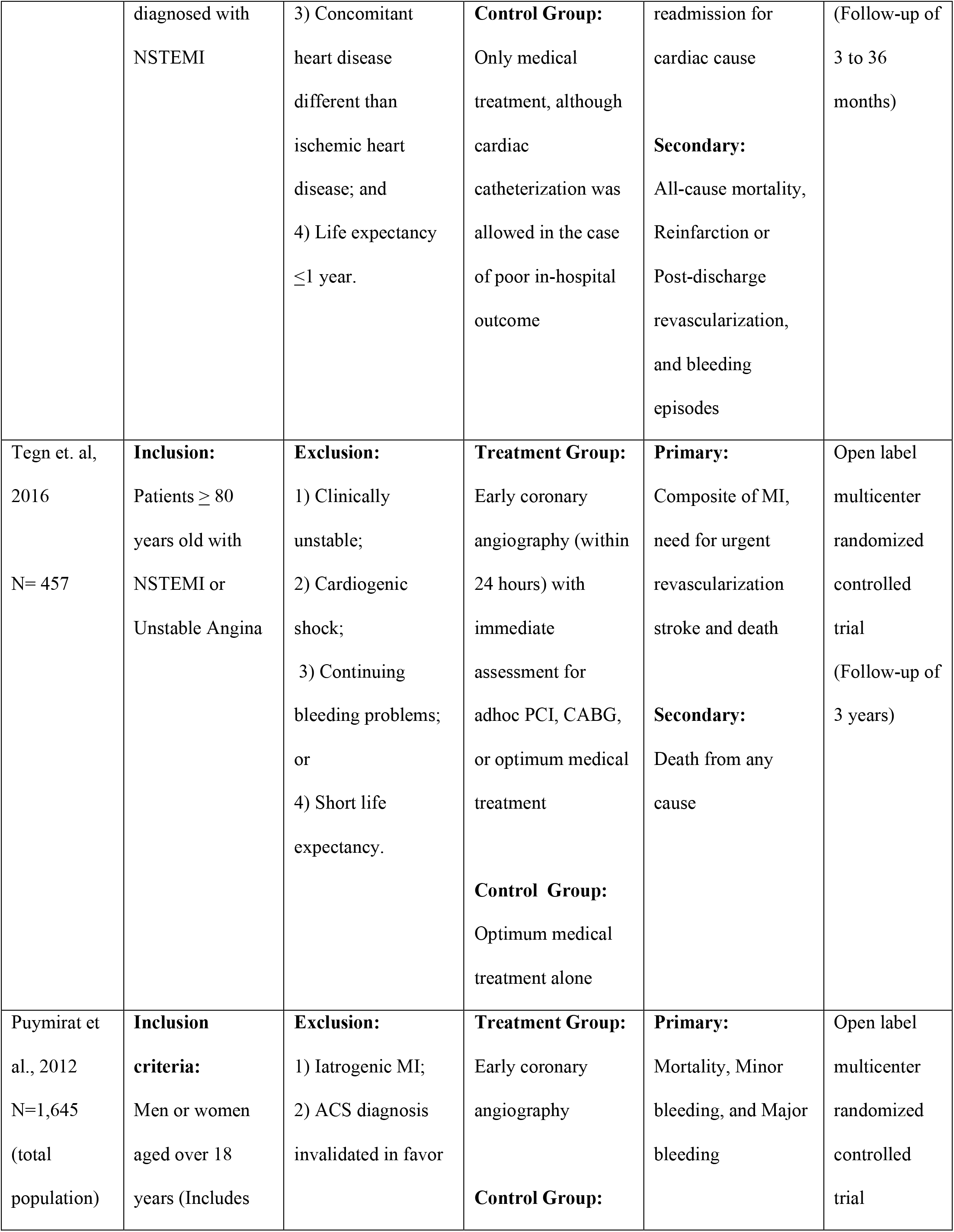
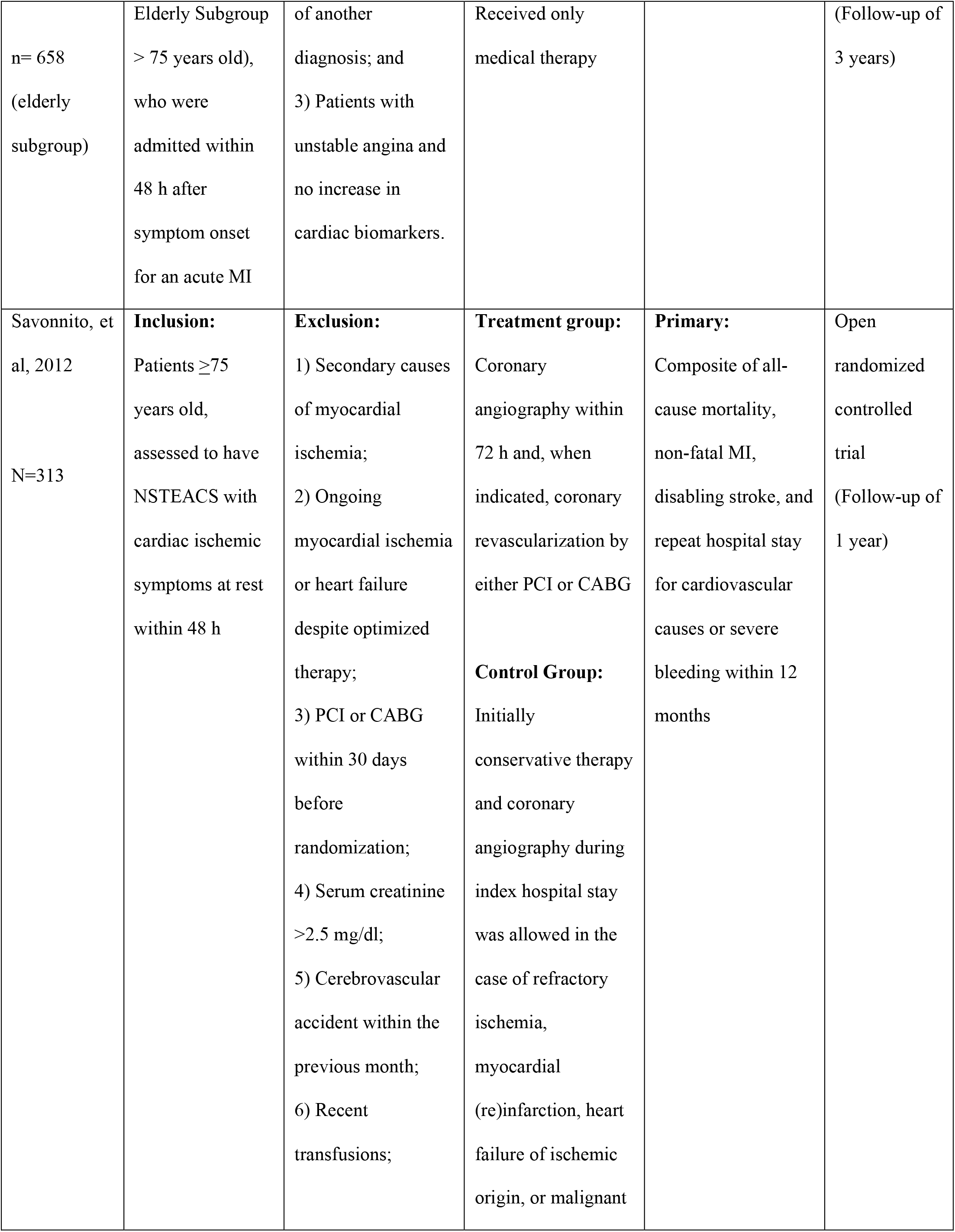
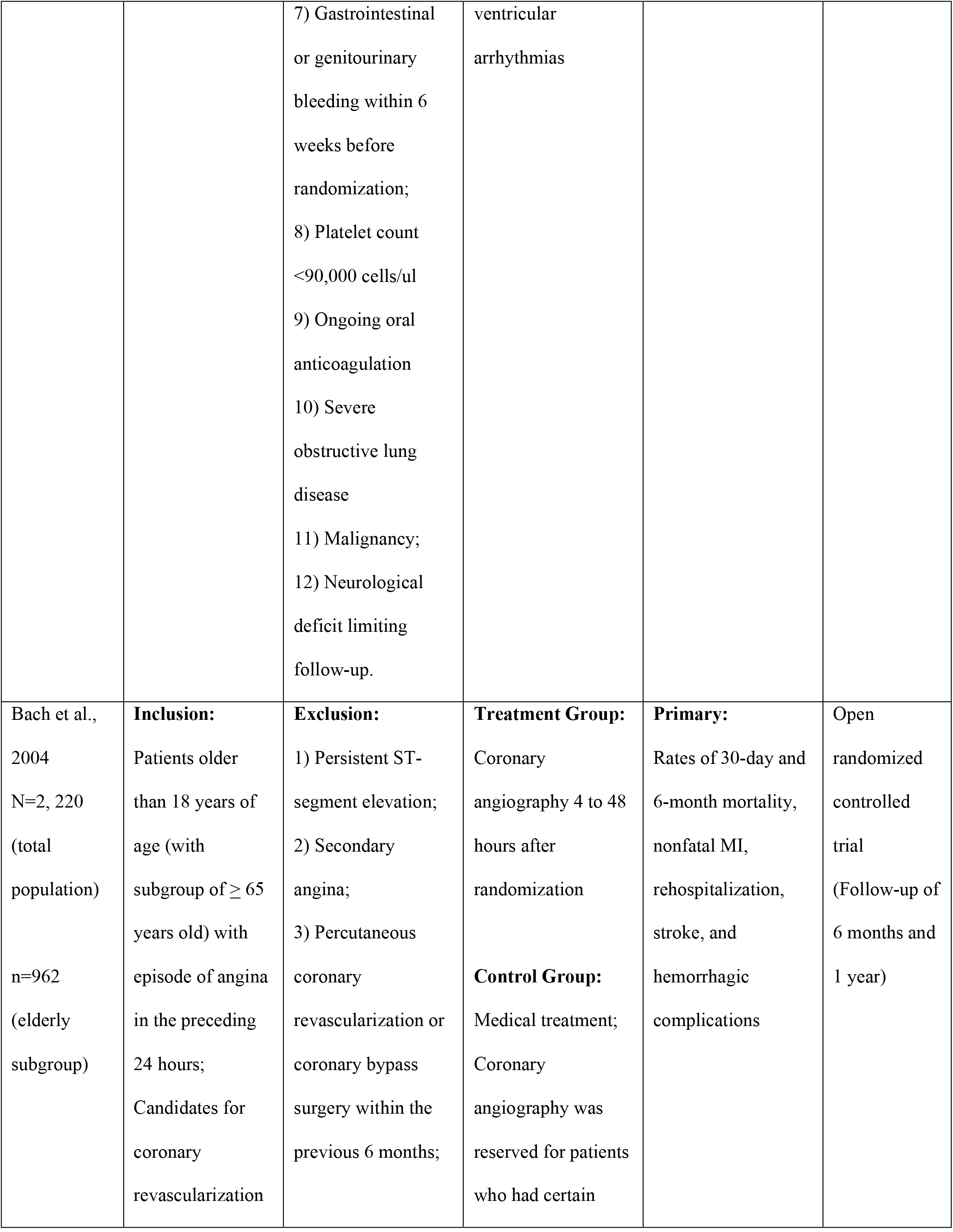
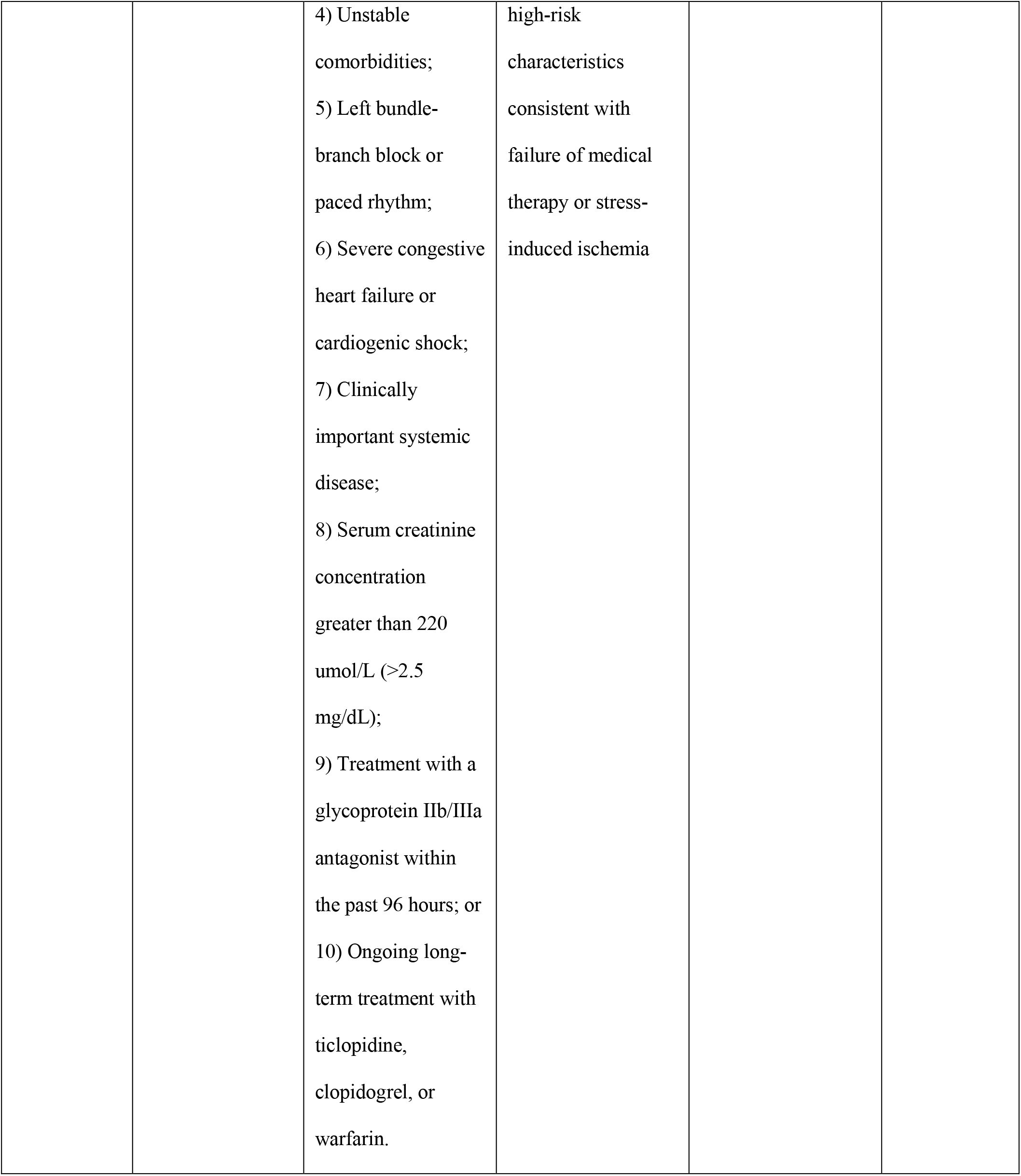
Characteristics of included trials

In the treatment arm, four trials specified the time to intervention (4-72 hours) ^10,12,13,14^. Only one study did not specify the time to intervention but only mentioned “during initial admission”.^11^ Two out of the five trials included CABG as part of the intervention when indicated.^12,13^ In the control group all the trials used standard medical treatment.^10-14^

All trials assessed the outcome of all-cause mortality. All trials except one reported the outcome of myocardial infarction.^11^ All trials except two assessed the outcome of stroke.^11,14^ The outcomes of revascularization were reported by all except by two studies.^10,11^ Lastly, the events of cardiovascular death and recurrent angina were assessed only by one study.^13^

The Cochrane collaboration tool was used to assess the risk of bias. The random sequence generation, allocation concealment, incomplete outcome data, blinding of participants and personnel, blinding of outcome assessment, and intention-to-treat analysis were evaluated for each trial. All included trials were assessed to have low risk for bias (Table 2).

**Table 2.** Quality assessment table.

| **Study ID** | **Method of Random Sequence Generation (Selection Bias)** | **Method of Allocation Concealment (Selection Bias)** | **Incomplete Outcome Data/Loss of participants to follow up (Attrition Bias)** | **Blinding of Participants and Personnel (Performance Bias)** | **Blinding of Outcome Assessment (Detection Bias)** | **Selective Reporting/ Intention to treat analysis (Reporting Bias)** |
| --- | --- | --- | --- | --- | --- | --- |
| Sanchis et al., 2016 | Low Risk | Low Risk | Low Risk | Low Risk | Low Risk | Low Risk |
| Tegn et. Al, 2016 | Low Risk | Low Risk | Low Risk | Low Risk | Low Risk | Low Risk |
| Puymirat<br>et al.,<br>2012 | Low Risk | Low Risk | Low Risk | Low Risk | Low Risk | Low Risk |
| Savonnito,<br>et al, 2012 | Low Risk | Low Risk | Low Risk | Low Risk | Low Risk | Low Risk |
| Bach et<br>al., 2004 | Low Risk | Low Risk | Low Risk | Low Risk | Low Risk | Low Risk |

## V. RESULTS

### Effects of intervention on outcomes of interest

#### A. All-cause mortality

A total of 242 among 1338 (18 %) elderly patients with NSTEACS died in the Invasive Strategy Group; while 296 died among 1158 (26 %) patients in the Conservative Group (Figure 2). The pooled analysis of all-cause mortality showed statistically significant benefit of invasive over conservative strategy with an overall risk ratio of 0.63 (95% CI 0.55 to 0.72) but with significant heterogeneity (p value of 0.0001, I^2^ =84%).

**Figure 2.**
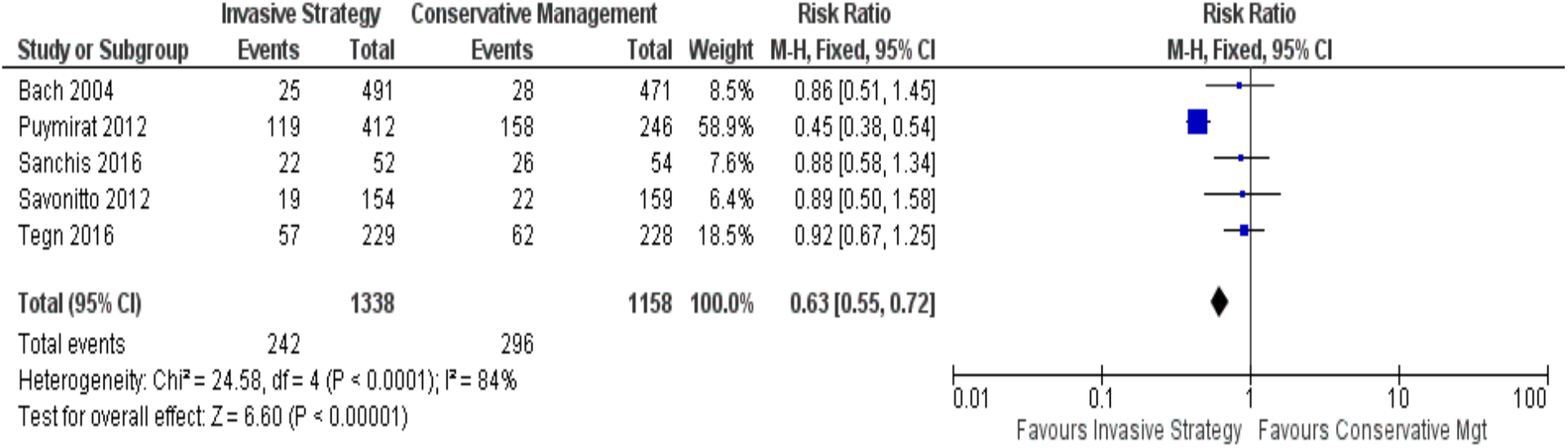
Comparison between invasive and conservative strategy with the outcome of all- cause mortality.

#### B. Myocardial infarction

In the Invasive Strategy Group, there were 89 events of MI among a total of 926 (10 %) patients; while there were 142 among 912 (16 %) patients in the Conservative Group (Figure 3). The pooled analysis showed that invasive strategy is beneficial over conservative treatment in preventing MI with an overall risk ratio of 0.62 (95% CI 0.49 to 0.79) but with significant heterogeneity (p value of 0.0001, I^2^ = 63%).

**Figure 3.**
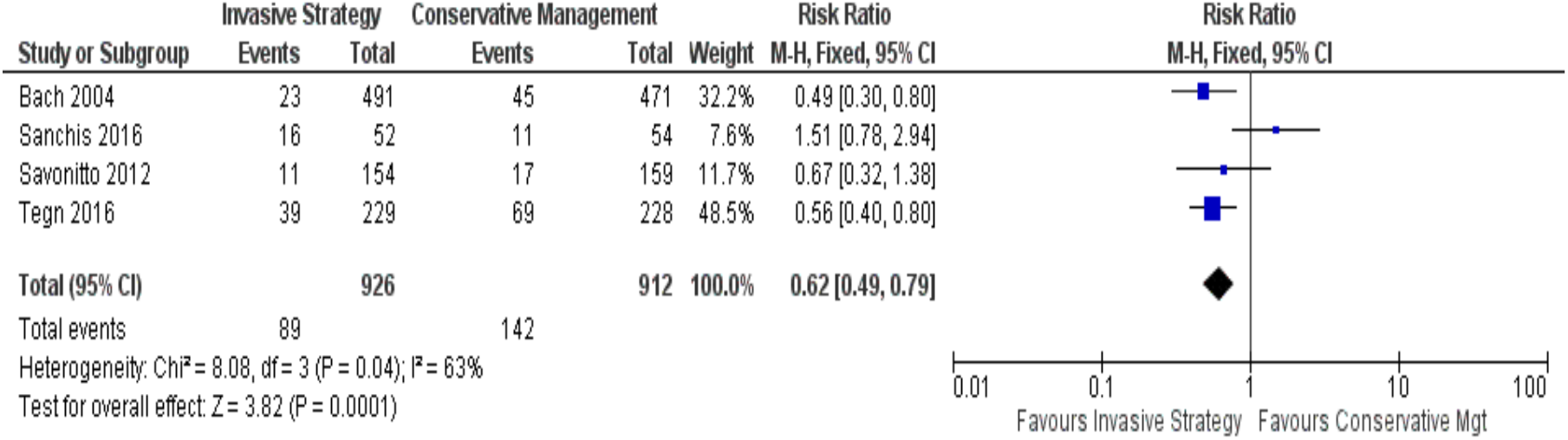
Comparison between invasive and conservative strategy with the outcome of myocardial infarction.

#### C. Stroke

Among the five trials, Savonitto et al. (2012), Tegn (2016), and Bach (2004) reported the outcomes of stroke (Figure 4). In the Invasive Strategy Group, there were 13 events of stroke among 874 (2%) patients; while there were 24 among 858 (3%) patients in the Conservative Group. The pooled analysis showed that early invasive strategy was favored over conservative treatment in preventing stroke but no statistically significant benefit with overall risk ratio of 0.53 (95% CI 0.27- 1.03, I^2^ =0%).

**Figure 4.**
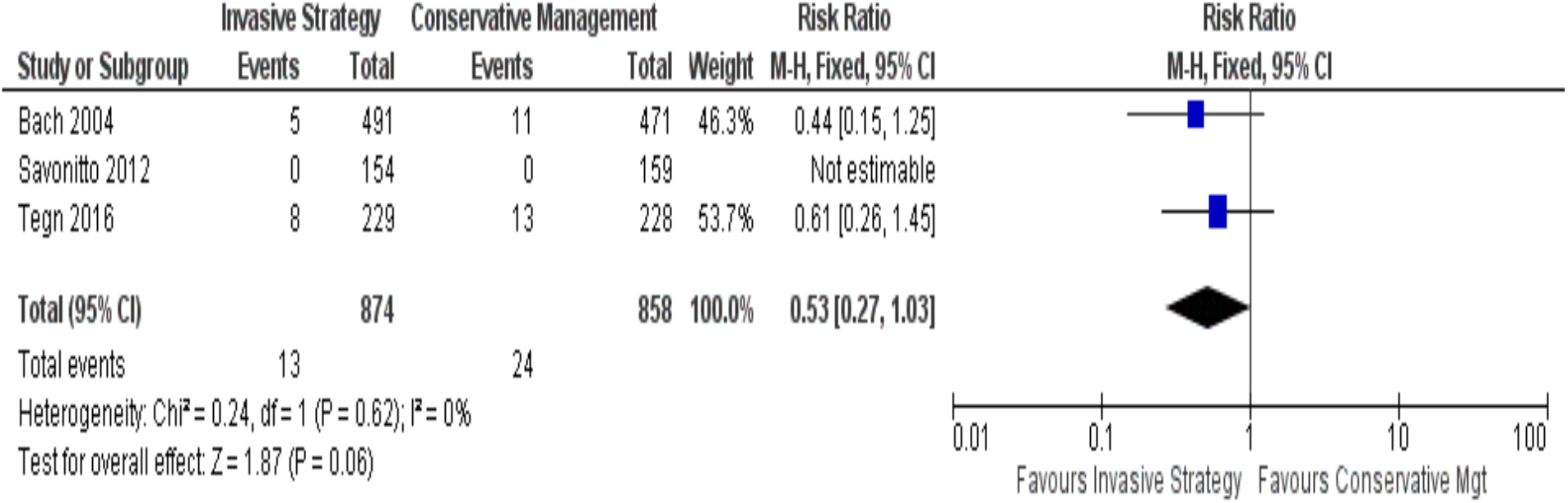
Comparison between invasive and conservative strategy with the outcome of stroke.

#### D. Need for revascularization

In elderly patients with NSTEACS, there were a total of 10 patients among 435 (2%) who needed revascularization in the Invasive Group while there were 34 patients among 441 (8%) in the Conservative Group (Figure 5). The pooled analysis for need for revascularization showed statistically significant benefit with an overall risk ratio of 0.31 (95% CI 0.16 to 0.61) with no significant heterogeneity (p value of 0.0006, I^2^ =0%).

**Figure 5.**
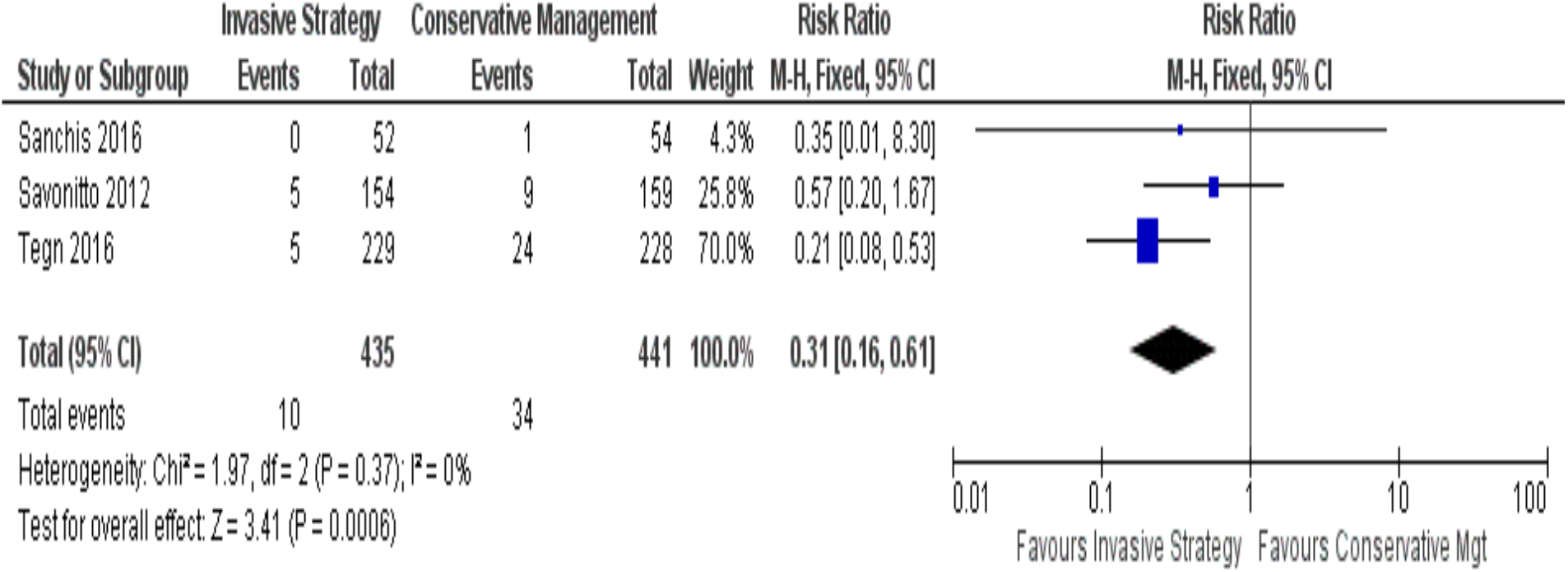
Comparison between invasive and conservative strategy with the outcome of need for revascularization.

#### E. Outcomes for cardiovascular mortality and recurrent angina

Among the five trials, only one trial assessed the outcomes of cardiovascular mortality and recurrent angina.^13^ The cardiovascular mortality incidence in the invasive versus the control group was 10% and 11 %, respectively, showing a non-statistically significant benefit of invasive over conservative treatment (RR 0.87, 95% CI, 0.49-1.56, p=0.65). Likewise, an invasive strategy showed a non-statistically significant benefit over conservative treatment in reducing recurrent angina (RR 0.81, 95% CI 0.45–1.46, p=0.49).

## VI. DISCUSSION

Meta-analysis of data from the five trials included in this study showed that an early invasive strategy appears to be beneficial in suitable elderly patients ≥ 65 years old with NSTEACS. There was significantly less need for revascularization in the invasive strategy group compared to the conservative treatment group. This finding implies that more patients in the conservative group clinically worsened during their course in the ward, requiring revascularization. It is also possible that early anatomic definition of the diseased coronaries may help the attending physician optimize an appropriate evidence-based management of the patient. The studies that evaluated the outcomes of revascularization stated that the indications for revascularization in the conservative group were: positive pre-discharge stress test, poor in-hospital outcomes, recurrent ischemia, reinfarction, malignant ventricular arrhythmias, refractory angina, and heart failure.^12-14^ Some patients who subsequently required revascularization could have probably been better off with an early invasive approach.

For the outcomes of death and MI, an invasive strategy showed a statistically significant benefit over conservative treatment but with significant heterogeneity. The possible sources of heterogeneity for the outcomes of death and MI may be the small number of events and sample sizes. In two studies, the elderly population was just a subgroup analysis of the total population.^10-11^ Hence, the population in the subgroup analysis may not be powered enough to detect the differences in the intervention and outcomes of interest. Furthermore, there were differences in age cutoffs and follow-up period. Two studies had age cutoffs of 75 years^11,13^ while the other three studies had age cutoffs of 65, 70, and 80 years.^10,12,14^ Possible clinical differences in outcomes may exist in these age brackets of the elderly population. In terms of follow-up periods, two studies had follow-up of 3 years^11,12^; one had follow-up period of 3 months to 3 years^14^; one had follow-up of 1 year^13^; while one had follow-up of 6 months and 1 year^10^. However, despite the heterogeneity, data from these studies clustered on the direction towards benefit favoring invasive over conservative strategy.

In the reduction of stroke, invasive strategy showed benefit over conservative treatment but this was not statistically significant. The outcomes for cardiovascular mortality and recurrent angina were assessed only in one study^13^, which showed also a non-statistically significant benefit of invasive strategy over conservative treatment among elderly NSTEACS patients.

Overall, this study does not support the relatively conservative tendency when dealing with elderly patients with NSTEACS in real-life clinical setting. The elderly population is considered a high-risk group wherein more than half the mortality in NSTEACS occur^5^ and a more aggressive approach in suitable patients may be more appropriate and beneficial. Among people who die of ischemic heart disease, 83% were >65 years of age.^1^ This mortality rate is expected to increase in the forthcoming decades due to improving life expectancy of the elderly. Age is one of the most important predictors of risk in NSTEACS. Each 10-year increase in age results in a 75% increase in hospital mortality in ACS patients.^15^ Despite the relatively higher risk in this age group, elderly ACS patients are under-represented in clinical trials such that subjects older than 75 years of age account for less than 10%, and those older than 85 years account for less than 2% of all NSTEACS subjects. ^7^ This highlights the need for more clinical trials and studies in this age group.

Data from the CRUSADE (Can Rapid Risk Stratification of Unstable Angina Patients Suppress Adverse Outcomes with Early Implementation of the American College of Cardiology/American Heart Association Guidelines) registry showed that NSTEMI patients aged ≥ 65 years who experienced an in-hospital major bleed had a 33% increased risk of 30-day mortality.^16^ However, the advancement of equipment and technique has made PCI safer for even very elderly patients (≥ 90 years of age) with high success rates and declining major bleeding risk.^17^

## VII. SUMMARY AND CONCLUSION

Results of this meta-analysis suggest some benefits with an early invasive strategy compared to a conservative treatment approach in the management of elderly patients with NSTEACS. There was a significantly lower rate of revascularization in the invasive strategy group compared to the conservative treatment group. A statistically significant benefit favoring invasive strategy was also noted in the reduction of death and myocardial infarction but with significant heterogeneity. These findings do not support the bias against early routine invasive intervention in the elderly group with NSTEACS.

Although an early invasive strategy may be favorable among elderly patients presenting with NSTEACS, the certainty of benefit versus risk still needs to be supported by larger clinical trials and registries with uniform age cutoff for elderly, particularly ≥ 65 years old, to provide high generalizability and statistical power. Current risk scoring systems such as the GRACE (Global Registry of Acute Coronary Events) Score, TIMI (Thrombolysis in Myocardial Infarction) Risk Score, and CRUSADE Bleeding Score are recommended in the initial evaluation of elderly patients presenting with NSTEACS. A special risk scoring may be developed to more accurately identify those who are suitable for an early invasive strategy, with an expected larger outcome and survival benefit.

## Data Availability

All data are available within this manuscript.

## VIII. ACKNOWLEDGEMENT

The authors would like to thank God; other consultants in the Section of Cardiology, Department of Internal Medicine, Manila Doctors Hospital; family, and friends for their support and patience in making this meta-analysis possible.

## XI. APPENDIX

### APPENDIX A PubMed Search Strategy

**APPENDIX A: PubMed Search Strategy**
| Recent queries<br>in pubmed |  |  |  |
| --- | --- | --- | --- |
| Search | Query | Items found | Time |
| #100 | Search (#42 AND #66 AND #99 AND #20) | 322 | 21:35:50 |
| #99 | Search (#92 OR #93 OR #94 OR #95 OR #96 OR #97 OR #98) | 3218012 | 21:26:41 |
| #98 | Search (#90 OR #91) | 50047 | 21:25:23 |
| #97 | Search (#88 OR #89) | 4189 | 21:25:01 |
| #96 | Search (#80 OR #81 OR #82 OR #83 OR #84 OR #85 OR #86 OR<br>#87 OR #88 OR #89) | 344281 | 21:24:32 |
| #95 | Search (#75 OR #76 OR #77) | 1549850 | 21:22:53 |
| #94 | Search (#72 OR #73 OR #74) | 831057 | 21:21:59 |
| #93 | Search (#69 OR #70 OR #71) | 1563389 | 21:21:19 |
| #92 | Search (#67 OR #68) | 13679 | 21:20:30 |
| #91 | Search revascularization | 50047 | 21:18:20 |
| #90 | Search need for revascularization | 3465 | 21:18:06 |
| #89 | Search recurrent chest pain | 2911 | 21:17:56 |
| #88 | Search recurrent angina | 2673 | 21:17:31 |
| #87 | Search cvd hemorrhage | 229 | 21:17:19 |
| #86 | Search cvd bleed | 210 | 21:17:04 |
| #85 | Search cvd infarct | 2332 | 21:16:47 |
| #84 | Search cerebral bleed | 72121 | 21:16:36 |
| #83 | Search cerebral hemorrhage | 53180 | 21:16:24 |
| #82 | Search cerebral infarct | 49028 | 21:16:10 |
| #81 | Search cerebrovascular event | 3648 | 21:16:00 |
| #80 | Search cerebrovascular accident | 275080 | 21:15:40 |
| #79 | Search cerebrovascular disease | 338376 | 21:15:16 |
| #78 | Search stroke | 272396 | 21:15:01 |
| #77 | Search heart attack | 229883 | 21:14:43 |
| #76 | Search MI | 1344629 | 21:14:23 |
| #75 | Search myocardial infarction | 223305 | 21:14:04 |
| #74 | Search cardiac death | 720781 | 21:13:45 |
| #73 | Search cardiovascular death | 95393 | 21:13:23 |
| #72 | Search cardiovascular mortality | 151179 | 21:13:03 |
| #71 | Search death | 720781 | 21:12:35 |
| #70 | Search mortality | 1044577 | 21:12:15 |
| #69 | Search all-cause mortality | 28210 | 21:11:59 |
| #68 | Search MACE | 6872 | 21:11:32 |
| #67 | Search major adverse cardiovascular events | 9103 | 21:09:35 |
| #66 | Search (#61 OR #62 OR #63 OR #64 OR #65) | 6249 | 21:08:38 |
| #65 | Search Invasive Therapy Conservative Therapy | 4294 | 21:07:35 |
| #64 | Search Invasive Treatment versus Conservative Treatment | 294 | 21:07:20 |
| #63 | Search Invasive Management versus Conservative Management | 183 | 21:07:07 |
| #62 | Search Invasive Strategy versus Conservative Strategy | 125 | 21:06:53 |
| #61 | Search (#59 AND #60) | 2471 | 21:06:15 |
| #60 | Search (#51 OR #52 OR #53 OR #54 OR #55 OR #56 OR #57 OR #58) | 125297 | 21:05:05 |
| #59 | Search (#43 OR #44 OR #45 OR #46 OR #49 OR #50) | 111701 | 21:03:38 |
| #58 | Search Optimal Medical Therapy | 42410 | 20:59:57 |
| #57 | Search Optimal Medical Management | 19244 | 20:59:42 |
| #56 | Search Optimal Medical Treatment | 48204 | 20:59:31 |
| #55 | Search Optimal Medical Strategy | 4920 | 20:58:27 |
| #54 | Search Conservative Therapy | 67332 | 20:58:09 |
| #53 | Search Conservative Treatment | 56611 | 20:57:58 |
| #52 | Search Conservative Management | 66213 | 20:57:43 |
| #51 | Search Conservative Strategy | 3336 | 20:57:30 |
| #50 | Search CABG | 15615 | 20:57:16 |
| #49 | Search Coronary Artery Bypass Graft | 64717 | 20:56:56 |
| #46 | Search PTCA | 41266 | 20:56:38 |
| #45 | Search Coronary Angioplasty | 46901 | 20:56:19 |
| #44 | Search Percutaneous Coronary Angioplasty | 21942 | 20:55:58 |
| #43 | Search Invasive Strategy | 9348 | 20:55:34 |
| #42 | Search (#40 AND #41) | 52265 | 20:53:53 |
| #41 | Search (#28 OR #29 OR #30 OR #31 OR #32 OR #33 OR #34 OR #35 OR #36 OR #37 OR #38 OR #39) | 118669 | 20:53:22 |
| #40 | Search (#21 OR #22 OR #23 OR #24 OR #25 OR #26 OR #27) | 7732231 | 20:51:38 |
| #39 | Search Q-wave myocardial infarction | 3366 | 20:49:40 |
| #38 | Search Q-wave MI | 757 | 20:49:29 |
| #37 | Search UA | 17483 | 20:49:13 |
| #36 | Search unstable angina | 17732 | 20:48:59 |
| #35 | Search ACS | 63075 | 20:48:41 |
| #34 | Search acute coronary syndrome | 25819 | 20:48:28 |
| #33 | Search non-Q wave myocardial infarction | 1631 | 20:48:10 |
| #32 | Search non-Q wave MI | 400 | 20:47:57 |
| #31 | Search NSTEMI | 2072 | 20:47:39 |
| #30 | Search non-st elevation myocardial infarction | 8832 | 20:47:25 |
| #29 | Search NSTEACS | 228 | 20:47:10 |
| #28 | Search non-st elevation acute coronary syndrome | 2893 | 20:46:51 |
| #27 | Search more than or equal to 65 years old | 3404034 | 20:46:33 |
| #26 | Search (65 years old and above) | 845 | 20:46:04 |
| #25 | Search super centenarian | 491 | 20:45:49 |
| #24 | Search centenarian | 752696 | 20:45:34 |
| #23 | Search Advanced age | 4671906 | 20:43:19 |
| #22 | Search old | 898369 | 20:42:56 |
| #21 | Search elderly | 4686863 | 20:42:37 |
| #20 | Search (#1 OR #2 OR #3 OR #4 OR #5 OR #6 OR #7 OR #8 OR #9 OR #10 OR #11 OR #12 OR #13 OR #14 OR #15 OR #16 OR #17 OR #18 OR #19) | 9528711 | 20:42:17 |
| #19 | Search (Not (animals [mh] NOT human [mh])) | 4353823 | 20:40:33 |
| #18 | Search volunteer* [tw] | 180971 | 20:40:13 |
| #17 | Search prospectiv* [tw] | 709909 | 20:40:00 |
| #16 | Search control* [tw] | 4598941 | 20:39:43 |
| #15 | Search prospective studies [mh] | 445018 | 20:39:27 |
| #14 | Search follow-up studies [mh] | 569279 | 20:39:03 |
| #13 | Search evaluation studies [mh] Schema: all | 0 | 20:38:42 |
| #12 | Search evaluation studies [mh] | 0 | 20:38:41 |
| #11 | Search comparative study [mh] Schema: all | 0 | 20:38:18 |
| #10 | Search comparative study [mh] | 0 | 20:38:18 |
| #9 | Search research design [mh:noexp] | 92025 | 20:38:05 |
| #8 | Search (((((singl* [tw] OR doubl* [tw] OR trebl* [tw] OR tripl* [tw] AND (mask* [tw] OR blind* [tw])) OR (placebos [mh] OR placebo* [tw] OR random* [tw])))) | 1225171 | 20:37:46 |
| #7 | Search ("clinical trial" [tw]) | 640470 | 20:37:25 |
| #6 | Search clinical trials [mh] | 303191 | 20:36:57 |
| #5 | Search clinical trial [pt] | 767368 | 20:36:46 |
| #4 | Search single-blind method | 39999 | 20:36:26 |
| #3 | Search double-blind method [mh] | 140472 | 20:36:09 |
| #2 | Search random allocation [mh] | 90997 | 20:35:54 |
| #1 | Search randomized controlled trials [mh] | 111611 | 20:35:19 |

### APPENDIX B Sample Data Extraction Template

**Sample Data Extraction Template**
|  |  |  |
| --- | --- | --- |
| Trial ID | Extractor | Year of publication |
| Title |  |  |
| Authors |  |  |
| Citation |  |  |

#### Participants

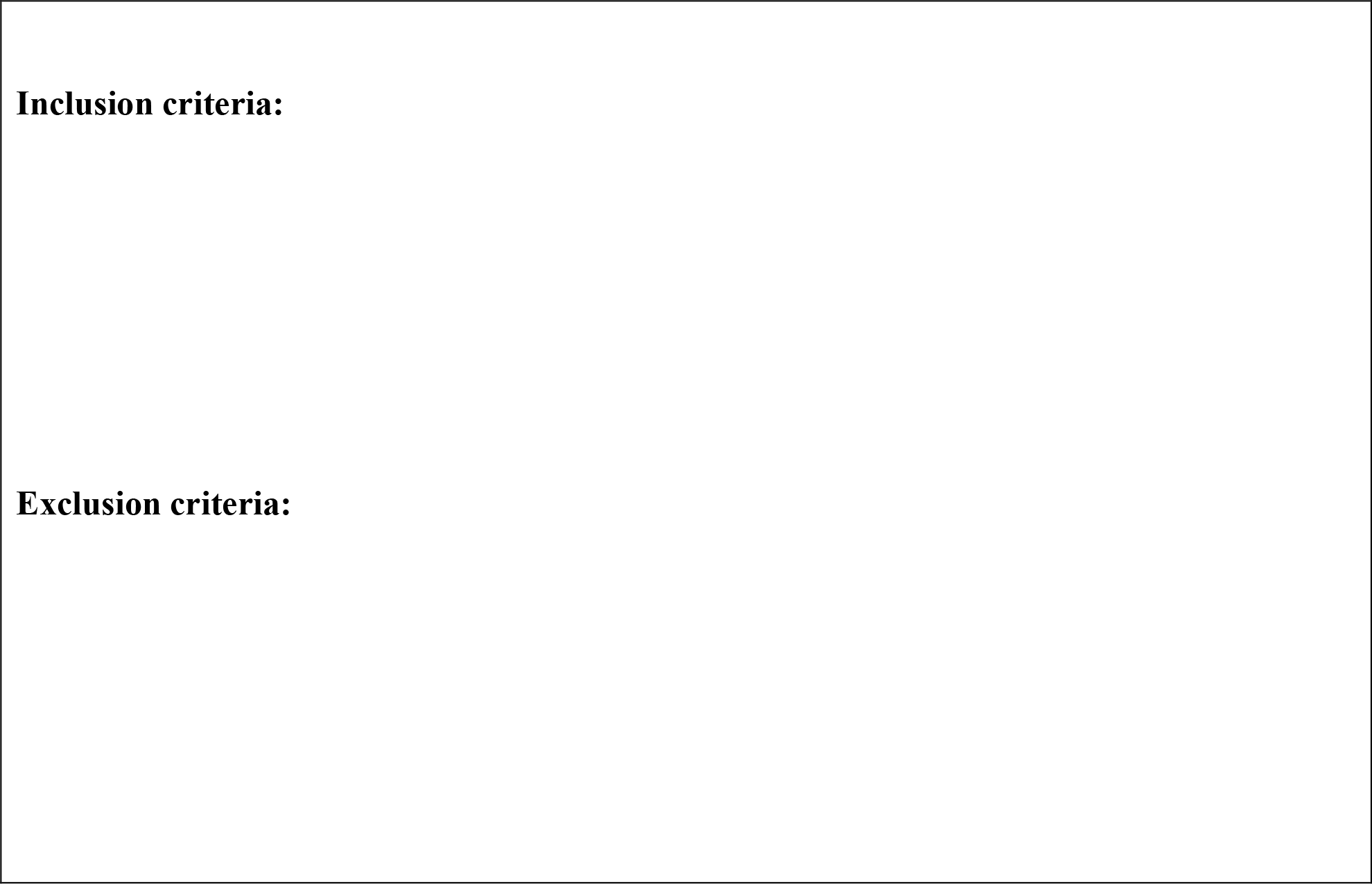

#### Intervention

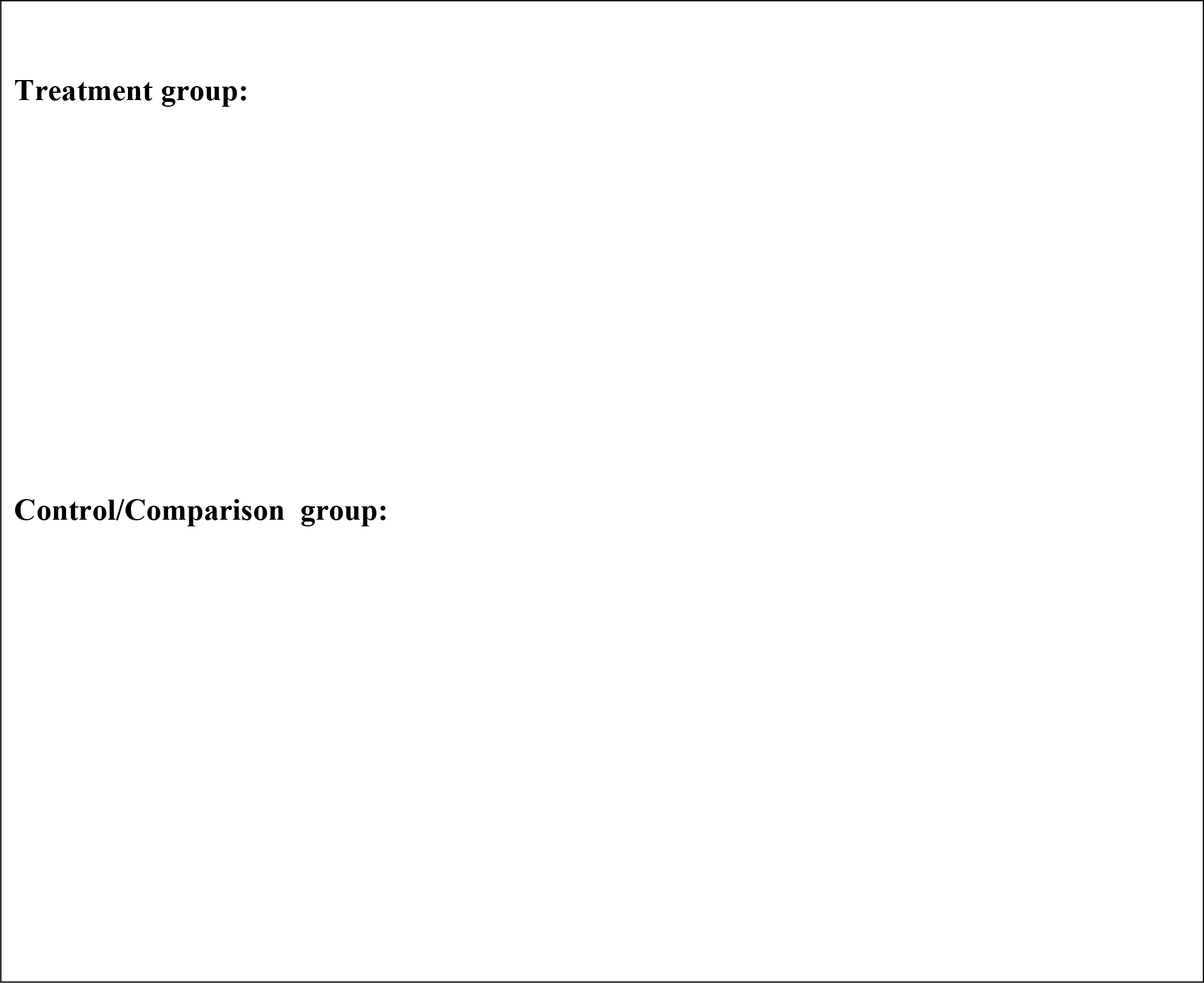

#### Method

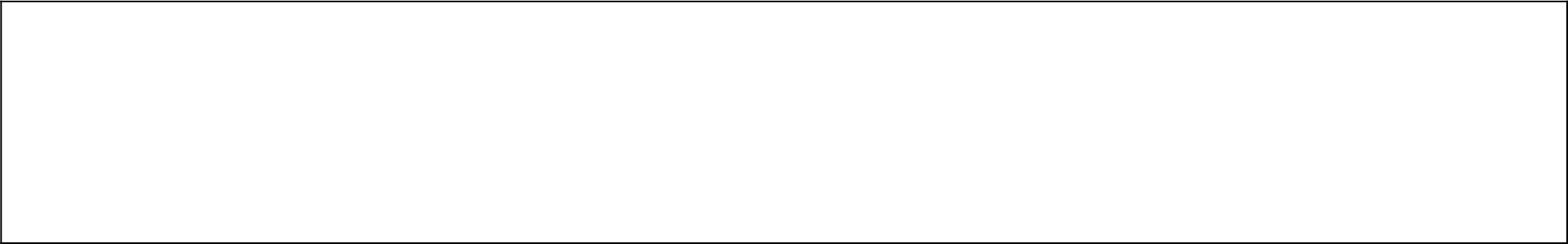

#### Quality assessment/Risk of Bias Table

**Quality assessment/ Risk of Bias Table**
| Domain | Judgement<br>Low Risk/ High Risk/ Unclear | Support for Judgement/<br>Description |
| --- | --- | --- |
| Method of Random<br>sequence<br>Generation<br>(Selection Bias) |  |  |
| Method of<br>allocation<br>Concealment<br>(Selection Bias) |  |  |
| Incomplete<br>Outcome Data/Loss<br>of participants to<br>follow up (Attrition<br>Bias) |  |  |
| Blinding of<br>Participants and<br>Personnel<br>(Performance Bias) |  |  |
| Blinding of<br>Outcome<br>Assessment<br>(Detection Bias) |  |  |
| Selective Reporting/<br>Intention to treat<br>analysis (Reporting<br>Bias) |  |  |
| Other Bias |  |  |

#### Outcomes

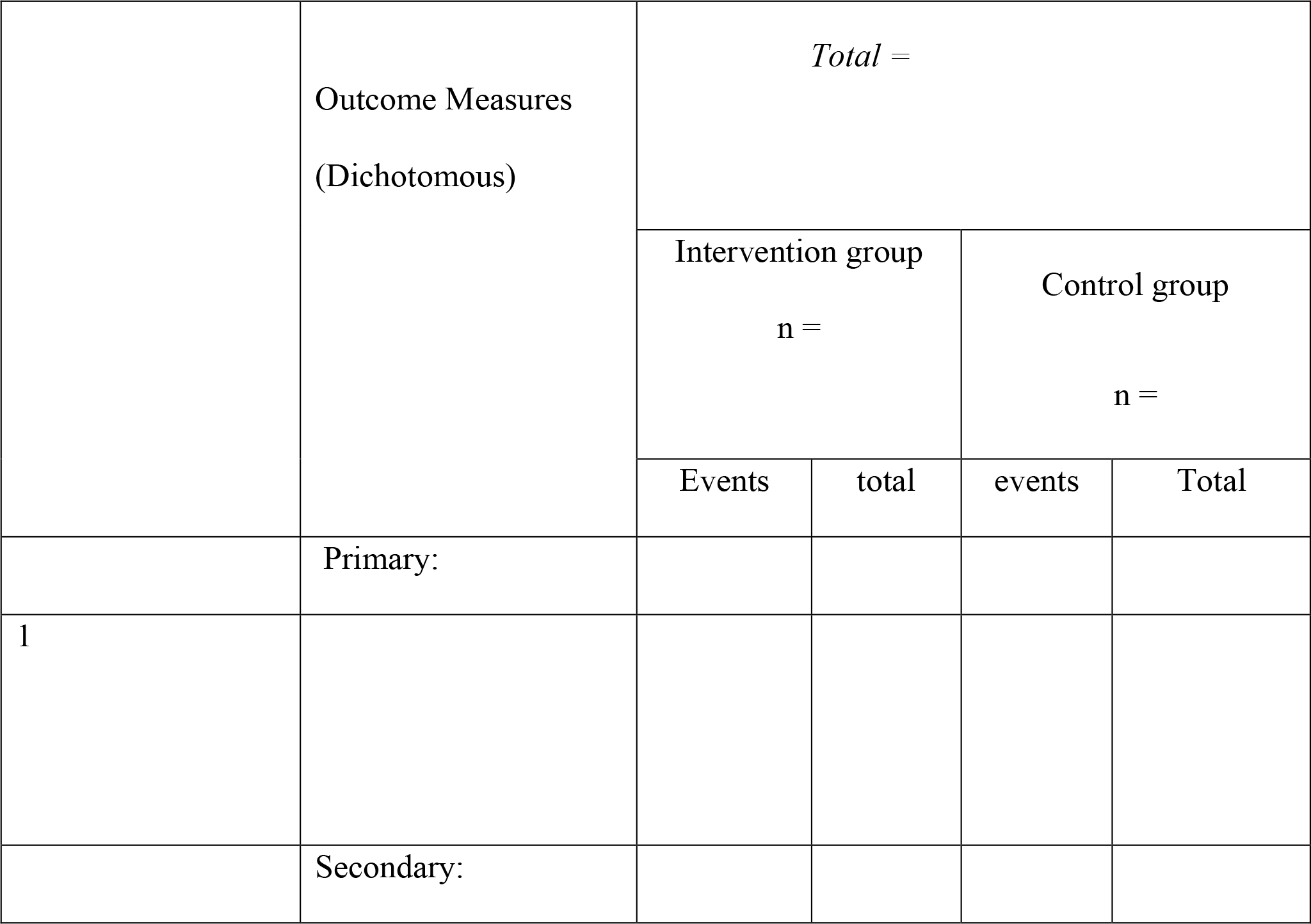
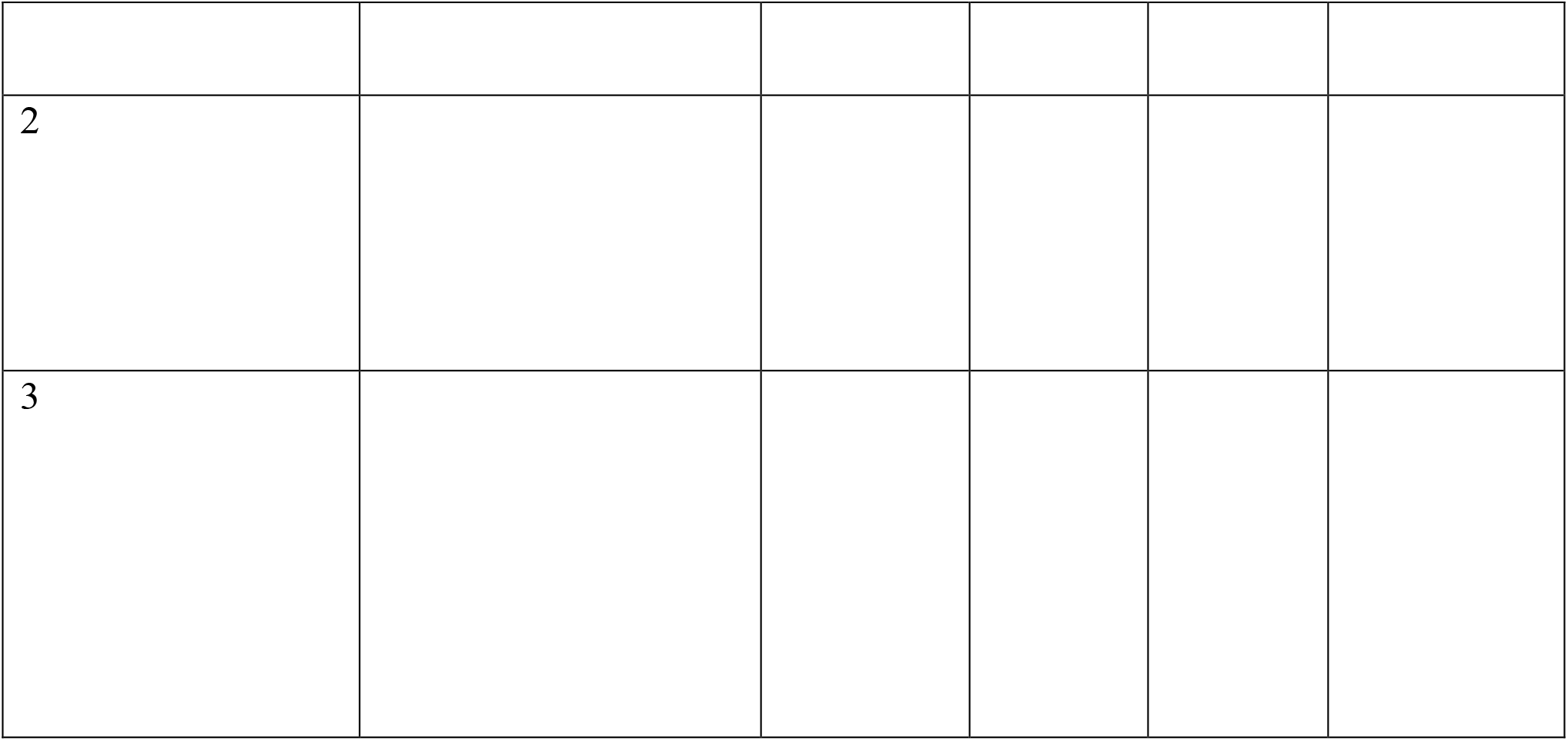

### APPENDIX C Summary of Results of the Five Included Randomized Controlled Trials

**Tegn et al**., **2016. After Eighty Study**

Invasive versus conservative strategy in patients aged 80 years or older with non-ST-elevation myocardial infarction or unstable angina pectoris (After Eighty study): an open-label randomised controlled trial

|  | Outcome Measures<br>(Dichotomous) | *Total = 457* |  |  |  |
| --- | --- | --- | --- | --- | --- |
|  |  | Intervention group<br>n = 229 |  | Control group<br>n = 228 |  |
|  |  | Events | Total | Events | Total |
| 1 | All-Cause Mortality | 57 | - | 62 | - |
| 2 | Cardiovascular Mortality | Not reported |  | Not reported |  |
| 3 | Myocardial infarction | 39 | - | 69 | - |
| 4 | Stroke | 8 | - | 13 | - |
| 5 | Recurrent angina | Not reported | - | Not reported | - |
| 6 | Need for revascularization | 5 | - | 24 | - |

**Sanchis et al**., **2016**.

Randomized comparison between the invasive and conservative strategies in comorbid elderly patients with non-ST elevation myocardial infarction

|  | Outcome Measures (Dichotomous) | *Total = 106* |  |  |  |
| --- | --- | --- | --- | --- | --- |
|  |  | Intervention group<br>n = 52 |  | Control group<br>n = 54 |  |
|  |  | Events | Total | Events | Total |
| 1 | All-Cause Mortality | 22 | - | 26 | - |
| 2 | Cardiovascular Mortality | Not reported | - | Not reported |  |
| 3 | Myocardial infarction | 16 | - | 11 | - |
| 4 | Stroke | Not reported | - | Not reported | - |
| 5 | Recurrent angina | Not reported | - | Not reported | - |
| 6 | Need for revascularization | 0 | - | 1 | - |

**Savonitto et al**., **2012**.

Early Aggressive Versus Initially Conservative Treatment in Elderly Patients With Non–ST-Segment Elevation Acute Coronary Syndrome

Early Aggressive Versus Initially Conservative Treatment in Elderly Patients With Non-ST-Segment Elevation Acute Coronary Syndrome
|  | Outcome Measures<br>(Dichotomous) | *Total = 313* |  |  |  |
| --- | --- | --- | --- | --- | --- |
|  |  | Intervention group<br>n = 154 |  | Control group<br>n = 159 |  |
|  |  | events | Total | Events | Total |
| 1 | All-Cause Mortality | 19 |  | 22 |  |
| 2 | Cardiovascular Mortality | 16 |  | 17 |  |
| 3 | Myocardial infarction | 11 |  | 17 |  |
| 4 | Stroke | 0 |  | 0 |  |
| 5 | Recurrent angina | 0 |  | 4 |  |
| 6 | Need for revascularization | 5 |  | 9 |  |

**Puymirat et al**., **2012. FAST-MI**

Use of Invasive Strategy in Non–ST-Segment Elevation Myocardial Infarction Is a Major Determinant of Improved Long-Term Survival

FAST-MI (French Registry of Acute Coronary Syndrome)

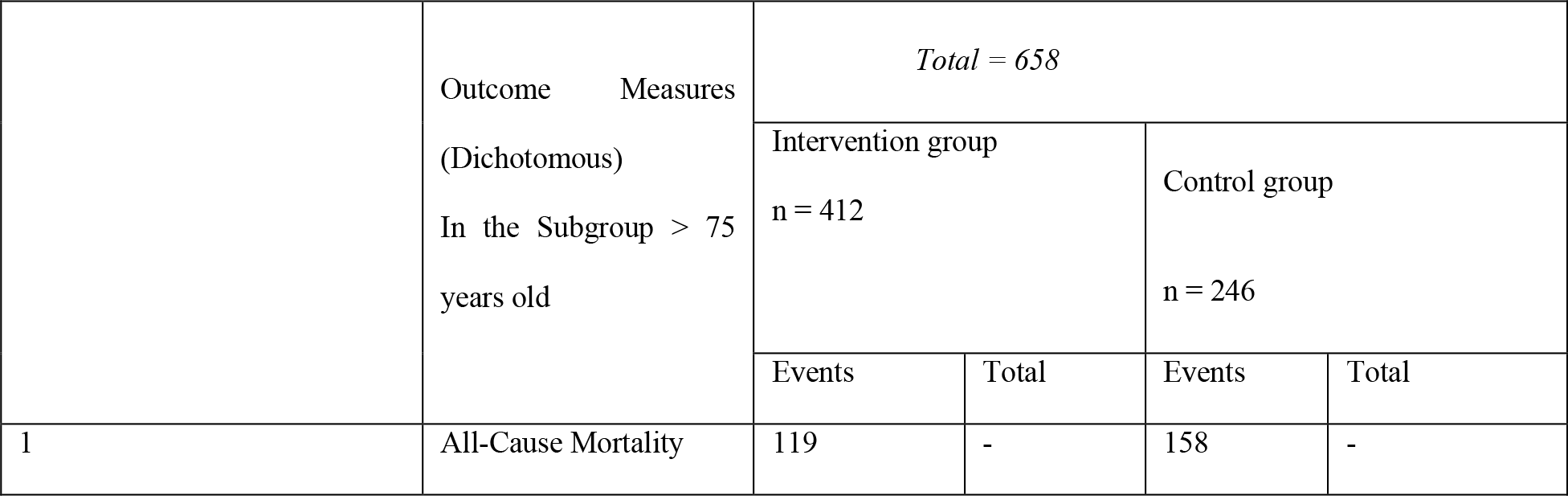
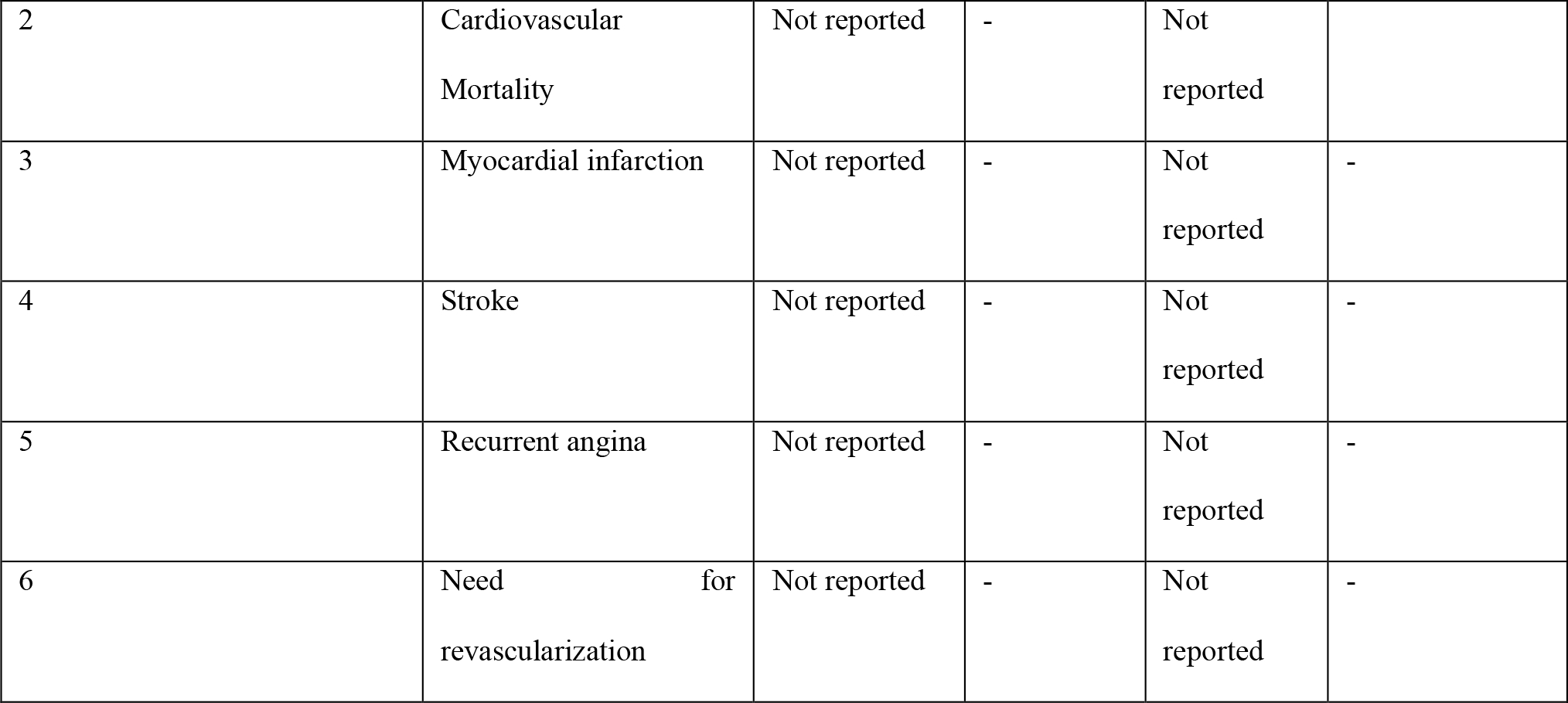

**Bach et al**., **2004**.

The Effect of Routine, Early Invasive Management on Outcome for Elderly Patients with Non–ST Segment Elevation Acute Coronary Syndromes

The Effect of Routine, Early Invasive Management on Outcome for Elderly Patients with Non-ST Segment Elevation Acute Coronary Syndromes
|  | Outcome Measures (Dichotomous) at 6 Months | *Total = 962* |  |  |  |
| --- | --- | --- | --- | --- | --- |
|  |  | Intervention group<br>n = 491 |  | Control group<br>n = 471 |  |
|  |  | Events | Total | Events | Total |
| 1 | All-Cause Mortality | 5.3 % (25) | - | 5.9% (28) | - |
| 2 | Cardiovascular Mortality | Not reported | - | Not reported |  |
| 3 | Myocardial infarction | 4.7 % (23) | - | 9.6 % (45) | - |
| 4 | Stroke | Not reported | - | Not reported | - |
| 5 | Recurrent angina | Not reported | - | Not reported | - |
| 6 | Need for revascularization | Not reported | - | Not reported | - |

### APPENDIX D Excluded Studies and Reasons for Exclusion

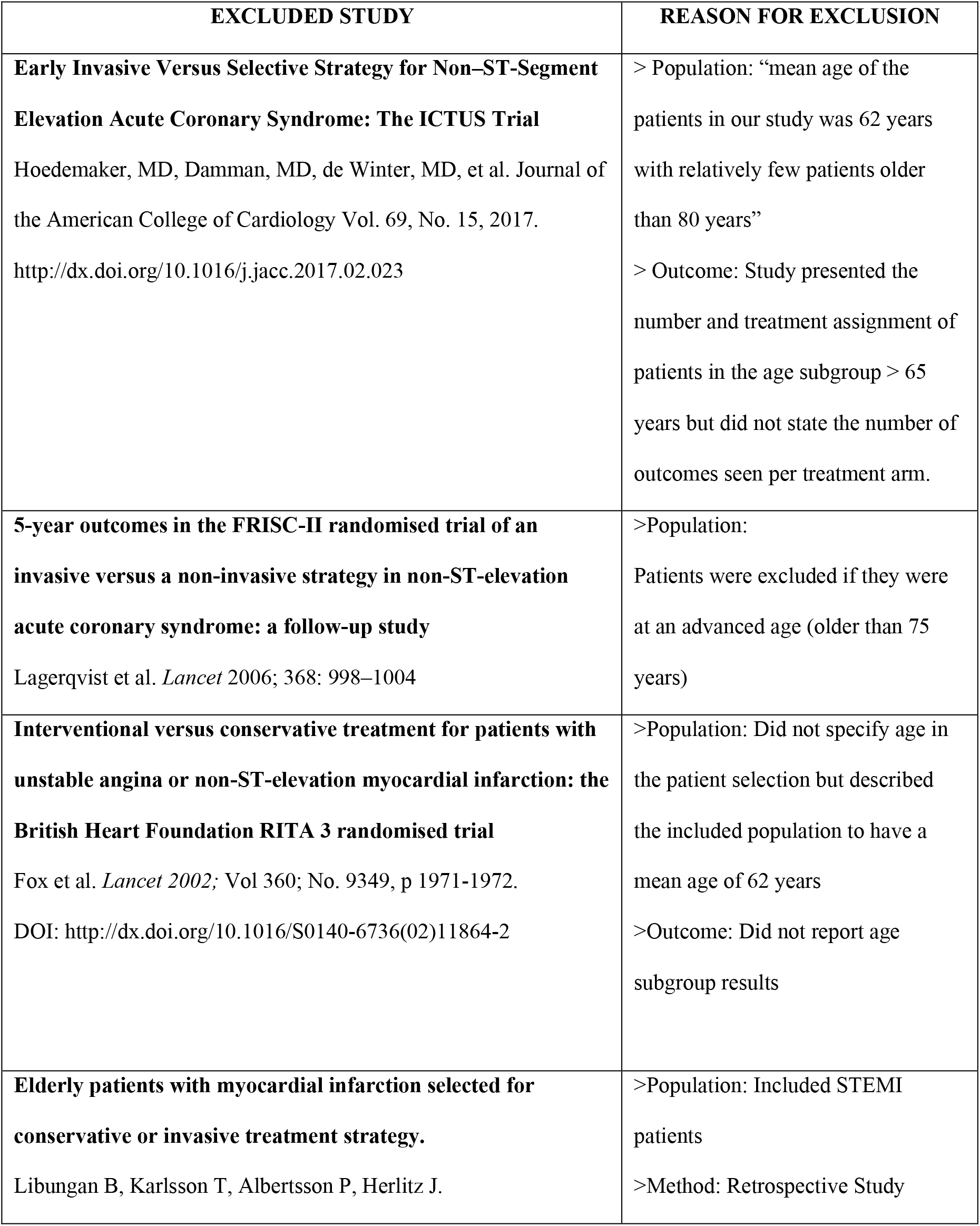
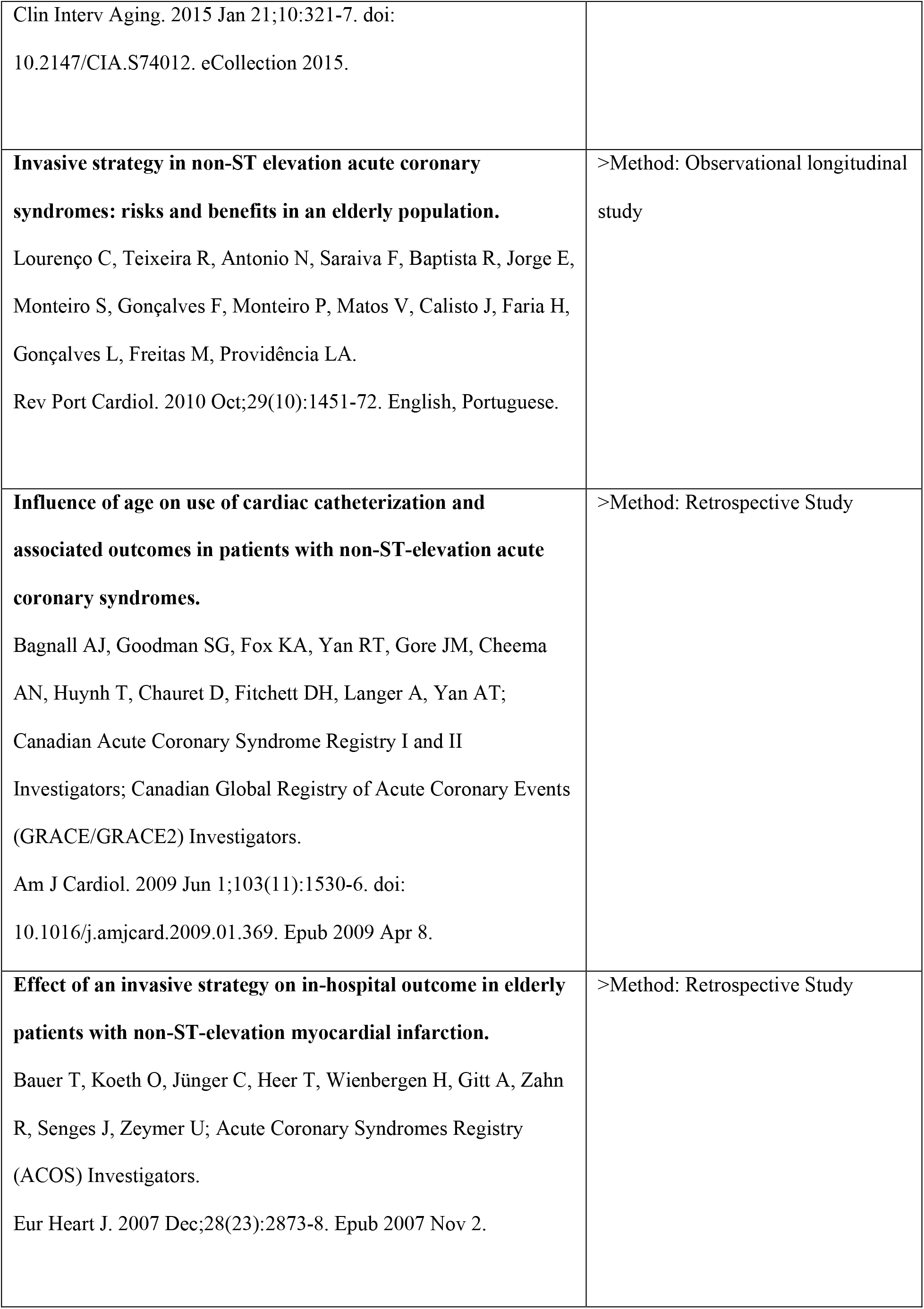
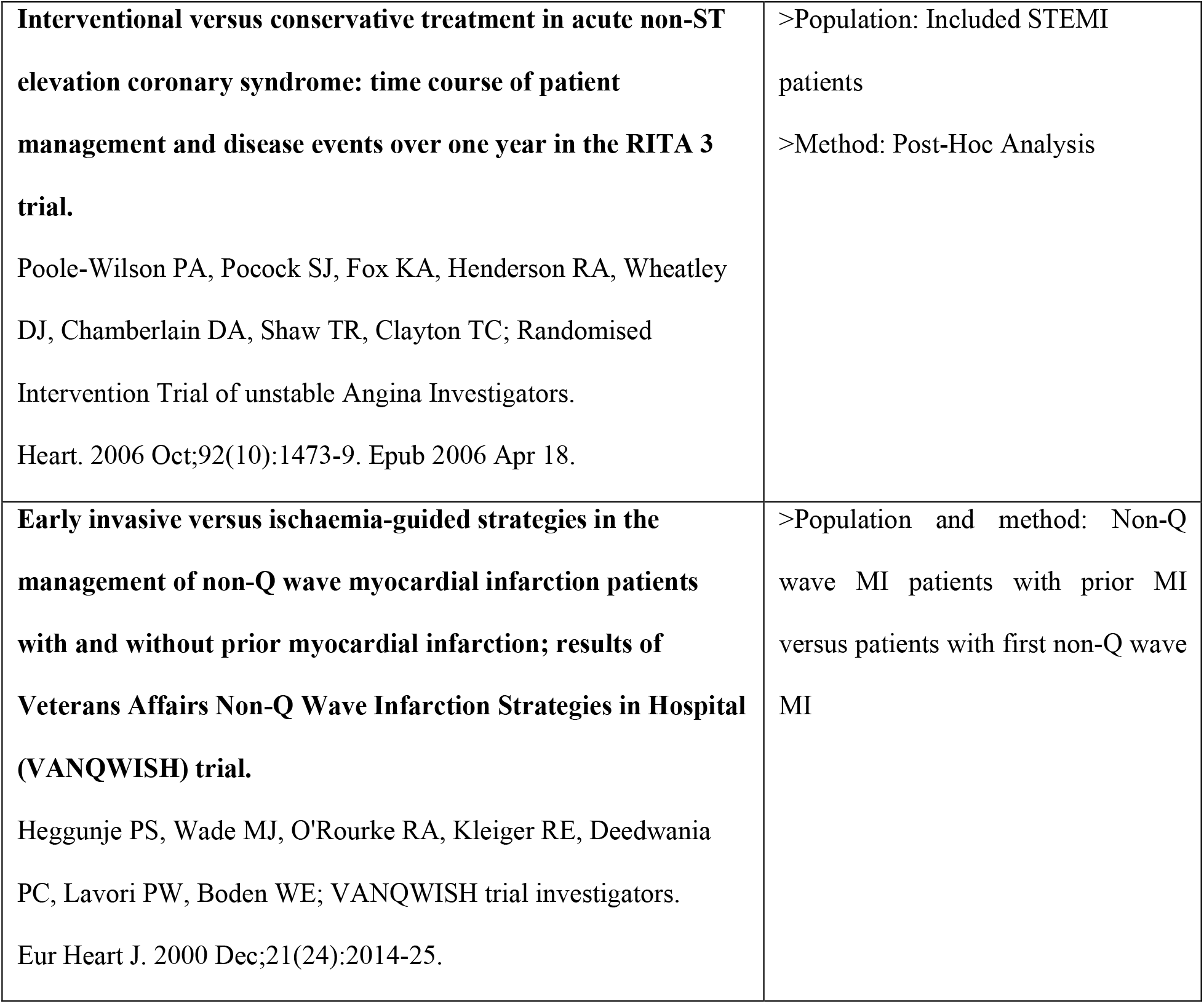

